# Customizing GPT-4 for clinical information retrieval from standard operating procedures

**DOI:** 10.1101/2024.06.24.24309221

**Authors:** Hannah Sophie Muti, Chiara Maria Lavinia Löffler, Marie-Elisabeth Leßmann, Esther Helene Stüker, Johanna Kirchberg, Malte von Bonin, Martin Kolditz, Dyke Ferber, Katharina Egger-Heidrich, Felix Merboth, Daniel E. Stange, Marius Distler, Jakob Nikolas Kather

## Abstract

**Background:** The increasing complexity of medical knowledge necessitates efficient and reliable information access systems in clinical settings. For quality purposes, most hospitals use standard operating procedures (SOPs) for information management and implementation of local treatment standards. However, in clinical routine, this information is not always easily accessible. Customized Large Language Models (LLMs) may offer a tailored solution, but need thorough evaluation prior to clinical implementation.

**Objective:** To customize an LLM to retrieve information from hospital-specific SOPs, to evaluate its accuracy for clinical use and to compare different prompting strategies and large language models.

**Methods:** We customized GPT-4 with a predefined system prompt and 10 SOPs from four departments at the University Hospital Dresden. The model’s performance was evaluated through 30 predefined clinical questions of varying degree of detail, which were assessed by five observers with different levels of medical expertise through simple and interactive question-and-answering (Q&A). We assessed answer completeness, correctness and sufficiency for clinical use and the impact of prompt design on model performance. Finally, we compared the performance of GPT-4 with Claude-3-opus.

**Results:** Interactive Q&A yielded the highest rate of completeness (80%), correctness (83%) and sufficiency (60%). Acceptance of the LLM’s answer was higher among early-career medical staff. Degree of detail of the question prompt influenced answer accuracy, with intermediate-detail prompts achieving the highest sufficiency rates. Comparing LLMs, Claude-3-opus outperformed GPT-4 in providing sufficient answers (70.0% vs. 36.7%) and required fewer iterations for satisfactory responses. Both models adhered to the system prompt more effectively in the self-coded pipeline than in the browser application. All observers showed discrepancies between correctness and accuracy of the answers, which rooted in the representation of information in the SOPs.

**Conclusion:** Interactively querying customized LLMs can enhance clinical information retrieval, though expert oversight remains essential to ensure a safe application of this technology. After broader evaluation and with basic knowledge in prompt engineering, customized LLMs can be an efficient, clinically applicable tool.

## Introduction

The field of medicine has experienced a substantial knowledge gain over the past century.[1] In healthcare systems across the globe, shortage of staff and aging populations make time a rare and valuable resource.[2] Digital transformation has been shown to increase workflow efficiency, patient satisfaction and physician well being.[3] Large Language Models (LLMs), a Natural Language Processing (NLP) based technology, have substantially evolved and been applied to clinical research.[4,5] Public interest was drawn to LLMs through Chat-GPT, an openly available LLM with a chat-based interface developed by OpenAI.[6] This made the technology accessible to the general public and raised a debate about its implications in healthcare. [7] LLMs have been shown to pass medical licensing exams[8,9], develop models for medical data analysis[10] or generate summaries of patient interactions and medical histories.[11] However, LLMs have also been criticized for delivering invalid information referred to as hallucinations, which could be harmful in a clinical setting.[10,12] With GPT-4, OpenAI introduced the option to customize LLMs through system instructions (system prompt) and submission of individual documents to the model.[13] This can be achieved through Retrieval Augmented Generation (RAG), where the documents and prompts are transformed into vector embeddings, over which a similarity search identifies relevant information within the embeddings.[14] This has been shown to achieve superior results compared to the use of a general LLM in medical use cases.[15]

In clinical routine, hospitals introduce best practice guidelines in the form of standard operating procedures (SOPs), which improves quality of care adapted to local treatment standards, resources or national guidelines.[16,17] Knowledge of a general LLM might not always concur with local standards, while a customized LLM can reduce the rate of hallucinations and the need for human intervention[18] when instructed to adhere to information provided in predefined documents.[19]

In clinical practice, fast access to reliable medical information is paramount to provide optimal patient care. Therefore, we created an interactive tool to access information form hospital-specific SOPs: We customized GPT-4 with 10 SOPs from four different departments of the University Hospital in Dresden, Germany and examined its ability to accurately answer 30 medical questions as they would occur in clinical routine of various degrees of detail. We evaluated accuracy within medical staff of various degrees of expertise and across different querying strategies. We assessed the impact of prompt design and non-text items in the SOPs on answer accuracy. Finally, we compared the browser-generated GPT with a self-coded RAG approach and benchmarked GPT-4 by OpenAI against Claude-3-opus by Anthropic. We show that a customized LLM can be used as an interactive information retrieval tool from an individual set of documents and we make our browser-generated GPTs and code publicly available. To our knowledge, this is the first study using an SOP-augmented LLM for medical information retrieval.

## Methods

### Ethics statement

The present study was conducted in accordance with the declaration of Helsinki. The study was approved by the ethics committee of the Dresden University of Technology under the reference number BO-EK-400092023. The study does not process patient-related data.

#### Data collection

We collected 10 SOPs from four different departments from the SOP database of the University Hospital Dresden, which were written in German. Four SOPs originated from the Department of Internal Medicine 1 (IM1), four SOPs originated from the Department of General Surgery (GS), one SOP originated from the Department of Clinical Infectiology (CIF) and one SOP originated from the Department of Pharmacology and Toxicology (PT). The 10 SOPs covered antibiotic treatment standards, COVID-19, opioids, neutropenia, CAR-T-Cell therapy, colorectal cancer, pancreatic cancer, oesophageal cancer, intestinal cleansing and intraperitoneal chemotherapy and were selected randomly to ensure diversity in length, content and complexity.

### LLM customization

With a pre-defined system prompt we used GPT-4 (accessed 16.03.2024) to customize a GPT based on our 10 SOPs, which we refer to as SOPHIA (SOP-based Hospital-specific Information Access). The system prompt can be found in Suppl. table 1. We redacted all SOPs to ensure author anonymity prior to uploading them to GPT-4. Web search, DALL-E use and the use of our uploaded data for internal purposes by OpenAI were denied. With the same protocol, we created a second GPT with a slightly modified system prompt (Suppl. table 1), which we named CARL (Clinical Assistant for Retrieval of Local information, accessed 21.03.2024). SOPHIA was prompted to be an assistant, CARL was prompted to be a doctor. Both system prompts contained instructions to answer questions asked by doctors querying the provided documents, to answer in a precise and professional way and to strictly adhere to the given information.

### Experimental design

Two physicians with clinical expertise from the departments of GS and IM1, respectively, created three questions of varying grades of detail and patient case vignettes per SOP. The questions can be found in Suppl. table 2 and were used for a question-and-answer analysis (Q&A) with the GPTs. The analysis was performed by 6 individuals of various degrees of professional experience: one medical student (observer 1), three residents, out of which two in IM1 and one in GS (observers 2-4) and two senior, board-certified physicians in IM1 and GS, respectively, who performed the Q&A according to their fields of expertise and who will be referred to as combined observer 5. (Fig. 1A) Observer 1, 3, 4 and 5 performed the analysis with SOPHIA, out of which observers 1, 3 and 5 conducted simple Q&A and observer 4 applied an interactive Q&A approach. Observer 2 performed the analysis with CARL. Each question was submitted once per observer, resulting in 30 questions per observer and 150 questions in total. Each observer submitted three “safety” questions querying for author names and information that was not contained in the SOPs, to verify adherence to the system prompt. These questions can be found in Suppl. Table 2.

**Figure 1.**
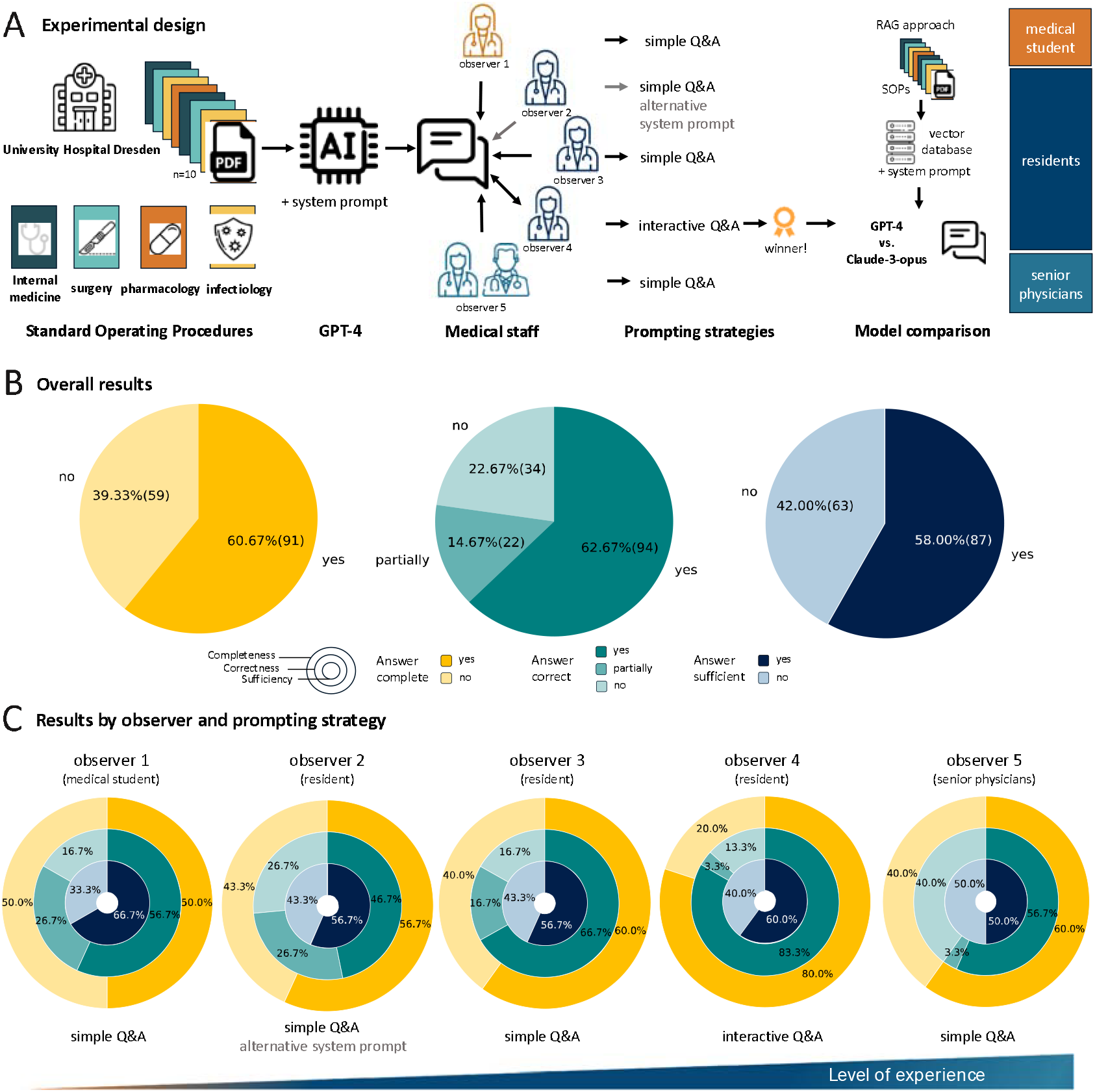
Experimental design and overall results. A: Experimental design: a publicly accessible GPT is customized with hospital-specific Standard Operating Procedures from different departments. 5 observers of various professional expertise query the GPT for hospital-specific information retrieval. The analysis is repeated within a RAG pipeline using the most successful experimental approach under comparison of two different Large Language models, GPT-4 and Claude-3-opus. B: Overall results covering completeness, correctness and sufficiency of the GPT’s answers for clinical use. C: Results by observer: completeness, correctness and sufficiency of the GPT’s answers for clinical use stratified by medical staff of various levels of professional expertise. All icons were obtained from https://www.flaticon.com/.

### Evaluation

Every observer individually interacted with the GPT and rated the given answers for completeness, correctness and sufficiency. Answers were defined as complete if every required detail was mentioned. Answers were defined as correct if every detail was reported according to the SOP, if the prompt requested a multitude of information of which only the majority was stated correctly (e.g. a list of drugs out of which one dosage interval was reported incorrectly), the answer was defined as partially correct. Answers were defined as sufficient if the observer found them suitable for use in a clinical routine setting. We furthermore designed the questions with different degrees of detail: Low detail questions targeted a broad section of an SOP and did not include patient- or case-specific information. Intermediate detail questions targeted a subset of a section of an SOP and contained one additional case-specific information detail. High detail questions targeted a subset of a section of an SOP and contained multiple case-specific details. We furthermore assessed the performance between genders in the patient case vignettes and SOP-specific characteristics like tables or flowcharts on the quality of the results with chi-squared tests.

### RAG analysis

In addition to OpenAI’s browser application, we constructed a pipeline using retrieval augmented generation (RAG) using the *LlamaIndex* framework (https://github.com/run-llama, first accessed 18.03.2024, last accessed 16.05.2024).[20] First, we generated vector embeddings of our PDF documents with *Chroma* (https://www.trychroma.com/, first accessed 18.03.2024, last accessed 16.05.2024) and stored them in a local vector storage.[21] Then, we created a chat engine to interact with the information in our vector storage under specification with our system prompt. Observer 3 repeated the analysis once with GPT-4 and once with Claude-3-opus via the OpenAI API and the Anthropic API, respectively. All code was written in *Python*. Hyperparameters are listed in Suppl. Table 3.

## Data and code availability

Our customized GPTs can be interacted with under https://chatgpt.com/g/g-MLkk5w66d-sophia (SOPHIA) and https://chat.openai.com/g/g-bFsNYtnu1-carl (CARL), respectively, with an OpenAI account. Due to local policy, we cannot make the SOP documents publicly available, however, the GPTs are finetuned with all SOPs used for this manuscript. All code is openly available under https://github.com/MutiHannah/SOPhia.

## Results

### Clinical-grade assessment of a customized LLM for information retrieval from standard operating procedures using GPT-4

We customized a GPT using OpenAI’s GPT-4 by supplying it with 10 SOPs in the form of PDF documents and assessed its ability to answer 30 predefined clinical-grade questions of various degrees of detail through observers of various degrees of experience. Six out of 10 SOPs contained plain text, six contained tables, two contained flowcharts, three contained pictures. The number of pages ranged from 1 to 11.(Table 1) Overall, our observers defined the GPT’s answers as complete in 60.67% of cases (vs. incomplete in 39.33% of cases), as correct in 62.67% of cases (vs. partially correct in 14.67% and incorrect in 22.67% of cases) and as sufficient in 58% of cases (vs. insufficient in 42% of cases). (Fig. 1B) We conclude that customized GPTs can retrieve information from a diverse set of SOPs, but that the overall approach can be optimized.

**Table 1.** SOP characteristics. Overview of all SOPs regarding the length in pages and the presence or absence of plain text, tables or flowcharts.

| SOP | number of pages | Contains plain text | Contains tables | Contains flowchart |
| --- | --- | --- | --- | --- |
| antibiotic treatment standards | 7 | no | yes | no |
| pancreatic cancer | 7 | yes | no | no |
| CAR-T-Cell therapy | 3 | no | yes | no |
| COVID-19 | 1 | yes | no | yes |
| intestinal cleansing | 1 | no | yes | no |
| neutropenia | 2 | yes | no | yes |
| intraperitoneal chemotherapy | 8 | yes | no | no |
| opioids | 1 | no | yes | no |
| oesophageal cancer | 11 | yes | yes | no |
| colorectal cancer | 1 | yes | yes | no |

### Interactive querying outperforms simple Q&A for clinical information retrieval

Furthermore, we compared the results of our Q&A with medical staff of various professional levels, hypothesizing that especially early-career medical staff would rate the GPT’s answers as helpful. The satisfaction with the given answers was indeed higher in early-career observers compared to the advanced professionals. (Fig. 1C) The student observer, despite reporting a lower proportion of complete and correct answers (50.0% and 56.7%), classified 66.7% of the answers as sufficient. (Fig. 1C, observer 1) Out of the early-career medical professionals, the best performance was achieved through the interactive approach (Fig. 1C, observer 4), which yielded the highest proportion of complete (80.0%) and correct (83.7%) answers. In 20 cases, one request was sufficient to obtain a suitable answer, one additional request was made in eight cases and 4 additional requests were made in 2 cases until a satisfactory answer was obtained. (Fig 2A) Senior physicians were most critical and classified 56% of the answers as correct and 50% as sufficient. (Fig. 1C, observer 5). The mean answering time with the GPT across all observers was 45 seconds with a standard deviation of 24 seconds compared to 144 seconds with a standard deviation of 133 seconds to find the respective information in our hospital’s knowledge database. (Fig. 2B and C) Finally, we assessed the GPT’s adherence to the system prompt through safety questions. As instructed, the GPT did not give away information beyond its set of SOPs. (Fig 2D) We conclude that with interactive Q&A, a customized GPT could be a clinically applicable information retrieval tool, but that professional expertise is irreplaceable to critically assess and contextualize given information. In addition, every observer reported discrepancies between the correctness and sufficiency of the given answers, so we administered further investigations.

**Figure 2.**
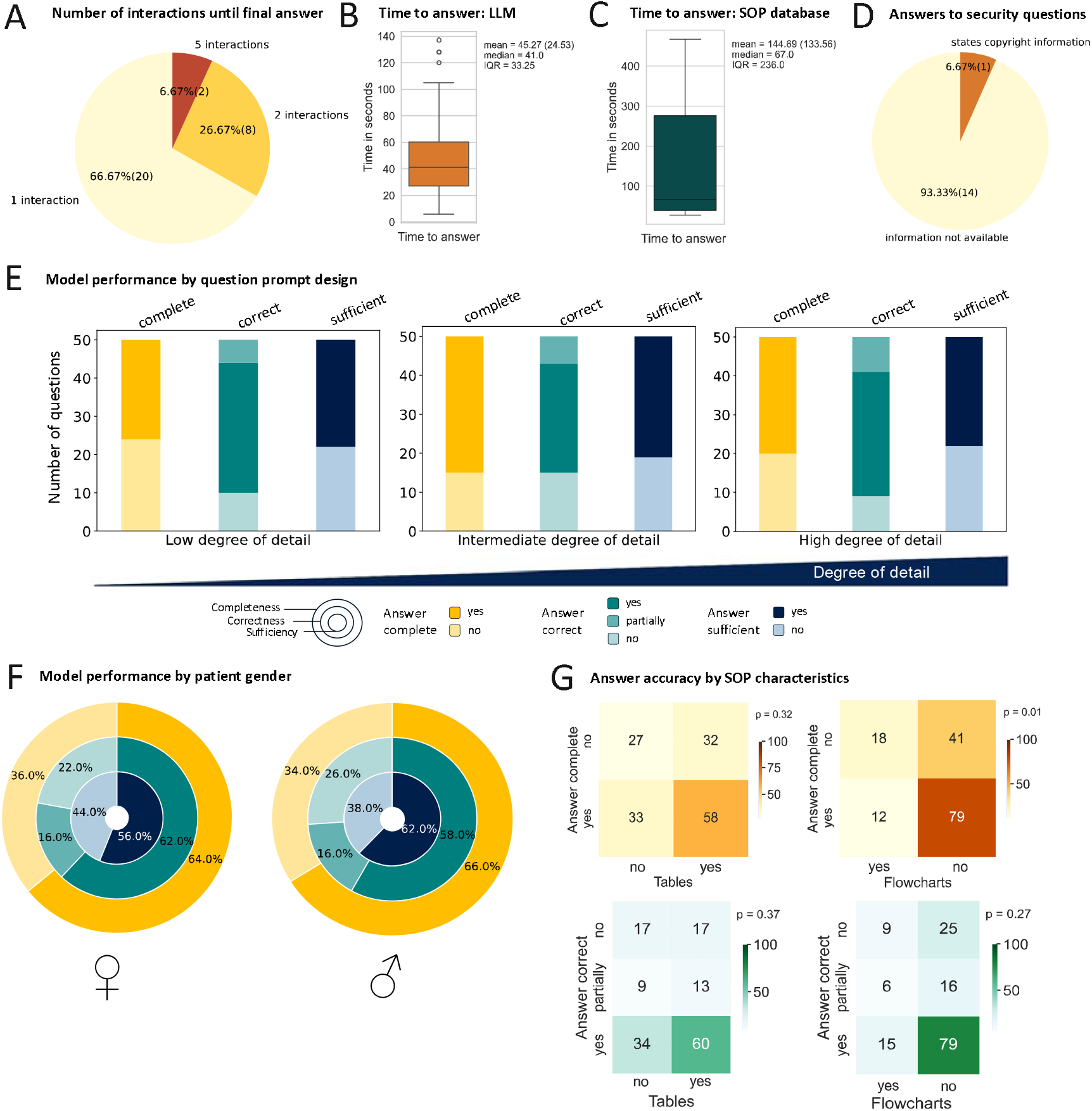
Results in respect to prompt design and SOP design. A: Pie chart showing the number of interactions needed to obtain a final answer through interactive querying for medical information through observer 4. B: Boxplot showing the time to retrieve information from an LLM. C: Boxplot showing the time to retrieve information from a classical knowledge database in a German university hospital. D: Assessment of the GPT’s answers to safety questions. E: Assessment of different prompting strategies to obtain answers: questions with low degree of detail contain no patient case-specific details and query for broad and unspecific information from a section of the SOPs. Questions with an intermediate degree of detail contain a patient-case specific aspect and query for a specific section of the SOPs. Questions with a high degree of detail contain more than one case-specific detail and query for precise information in a sub-section of the SOPs. F: Comparison of answer completeness, correctness and sufficiency by gender of the patient in the constructed case-vignettes. G Assessment of the completeness and correctness of the given answers by the presence or absence of tables or flowcharts in the given SOP through chi-squared tests.

### Prompt engineering optimizes answer quality

We hypothesized that the accuracy of GPT-generated answers might depend on prompt engineering or SOP-specific characteristics. To test this, we examined how question complexity affected answer quality.

Per SOP, we asked one highly, one intermediately and one non-detailed question. We found that the rate of complete and correct answers was highest with the intermediate and high detail prompts, while there was no notable difference between the sufficiencies of the answers according to question detail. (Fig. 2E) In German, unlike in English, all nouns, including the noun “patient”, have a gender. Hence, we analyzed model performance by gender of the patient in the given case-vignette, which was not the case. (Fig. 2F) Overall, answer sufficiency did not seem to depend on question design or patient gender, hence, we analyzed the impact of non plain text items on the completeness and correctness of the given answers. The presence of tables did not impact the GPT’s performance, whereas the presence of a flowchart seemed to reduce completeness (p = 0.01) without having any impact on the correctness of the answers (p = 0.27). (Fig. 2G) We conclude that prompt engineering is important for the quality of the answers and that the most suitable answers can be achieved through precise but not extensive prompts.

Moreover, prompt engineering, patient gender or the presence of non-text items in the SOP documents did not explain discrepancies between correct and sufficient answers.

### Claude-3-opus outperforms GPT-4

On the basis of our previous results, we chose the most successful prompting strategy, interactive Q&A, to compare our results which were generated through a browser-based application with a RAG system while comparing GPT-4 by OpenAI with Claude-3-opus by Anthropic. Using the same question prompts, the rate of correct answers was higher with GPT-4 than with Claude-3-opus (66.7% compared to 53.3%, Fig. 3A and C), whereas the rate of complete and sufficient answers was higher with Claude-3-opus than GPT-4 (66.7% vs. 43.3% and 70.0% vs. 36.7%, Fig. 3 C and A). In addition, the number of requests was higher with GPT-4 (Fig. 3B) than with Claude-3-opus (Fig. 3D). In seven cases, we were unable to obtain an answer through GPT-4 because the model claimed it was not supplied with the respective information (which was not the case) compared to Claude-3-opus, where this happened once (Fig. 3D). In the browser-based approach, this never occurred. (Fig. 2A) Given that the system prompt instructed the models to deny answers in case of an information deficit, self-coded pipelines seem to be less prone to giving faulty information than the browser application, while Claude-3-opus was more flexible in this setting.

**Figure 3.**
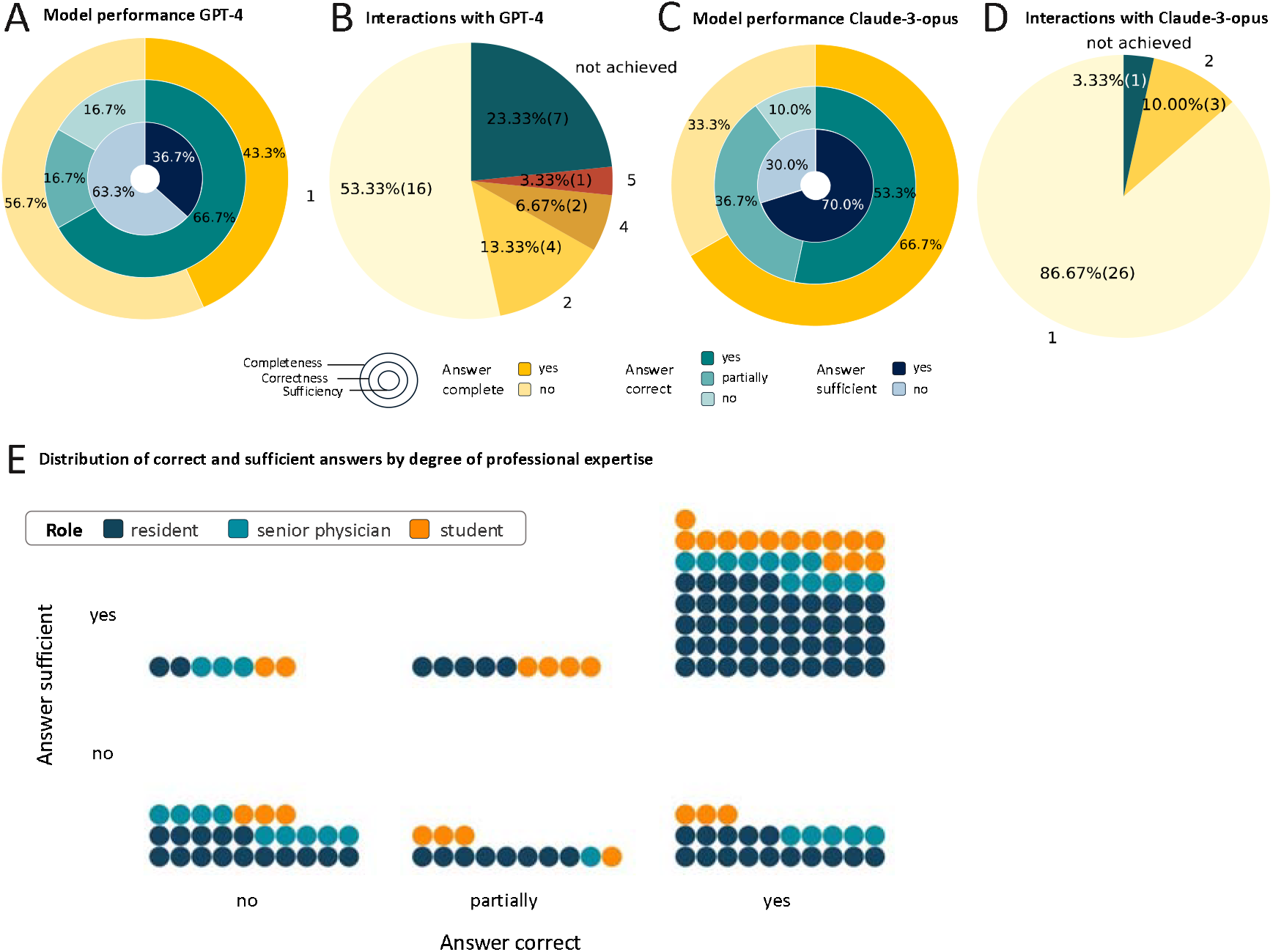
Comparison of GPT-4 and Claude-3-opus and distribution of answers perceived as correct vs. sufficient stratified by professional expertise of the observers. A Q&A results achieved with GPT-4. B Number of iterations needed with GPT-4. C Q&A results achieved with Claude-3-opus. D Number of iterations needed with Claude-3-opus. E Bubble chart showing the distribution of correct vs. sufficient answers stratified by professional expertise with each bubble representing one Q&A pair.

## Discussion

In the present study, we customized an LLM with SOPs from a German university hospital. We assessed the GPT’s performance in answering clinical-grade questions across observers from various professional levels and through different prompting strategies. We show that through interactive prompting, GPTs are suitable for streamlined information retrieval from preselected sources. According to our results, self-constructed GPTs are less prone to misinformation compared to pre-designed browser applications.

Truhn et al. argued that instead of single-shot Q&A, the true potential of LLMs lies in interactive reasoning[22], which our findings clearly corroborate.(Fig. 1C) However, discrepancies between correctness and sufficiency rates in our results indicate that an LLM’s performance is tied to the underlying sources of information.[23,24] According to our early-career observers, non-text information was better understandable in the original SOP than in the GPT-produced answer through human interpretation. However, information on further steps or when to consult senior professionals was not included in most SOPs but in the GPT’s answer, which was perceived as helpful. Hence, early-career staff can substantially profit from this technology, but is also substantially prone to misinformation through it, as previously discussed.[7] Correspondingly, we observed that senior physicians were more distinct in rating answers for correctness and sufficiency compared to junior observers. (Fig. 3C) Especially when senior expertise is unavailable, quick and easy access to professional information is invaluable for patient (and physician) wellbeing - standards save lives.[25] Notably, GPT-generated answers included statements on patient wellbeing and social situations, which only one SOP took into account. In line with this, LLMs have been described to outperform physicians in the expression of empathy before.[26,27] In line with ethical and practical considerations for the purpose of generative AI in medicine, with a tool like ours, clinical-grade information can be made available to hospital staff quickly and easily accessible.[28]

The idea that precisely asked questions improve answer quality accounts for human beings and LLMs equally. However, humans have a superior ability to derive information from context compared to LLMs.[29] For safe clinical use, basic prompt engineering skills will be paramount for optimal utilization of this technology and LLM-related education is expected to be integrated into medical workflows and education.[30]

Remarkably, all SOPs, prompts and answers were in German language. In line with previous findings, our results gave no evidence that language had any impact on our output quality.[31] As healthcare systems become more diverse and globalized[32], it would be highly relevant to leverage multilingual LLMs and assess prompting through non-native speakers.

### Limitations

Our study has several limitations. First and foremost, 10 SOPs are not representative of a hospital’s entire SOP database. After this initial proof-of-concept, our approach should be refined with a larger and more diverse set of documents, as described before.[19] Moreover, for economic purposes, the use of an open-source LLM like Llama should be evaluated in this use case.[33,34], In addition, incorrect answers can be harmful when searching for an adequate therapy for patients in potentially life-threatening conditions. However, in prior research LLMs outperformed human beings in medical information reproduction, the effects of this should be further analyzed.[35] In addition, there were discrepancies between correct and sufficient answers which based on our results cannot be explained through technical details. However, LLMs can uncover inconsistencies within a dataset and an LLM’s answer can only be as good as their source of information.[19] Lastly, we did not include nurses or physiotherapists into our experimental setup. In follow-up studies, every group of healthcare professionals should be integrated into method development in order to tailor applications to the users’ needs and to increase acceptance of this technology.[36,37]

### Outlook

LLM applications in healthcare are a dynamic area of research. In the recent months alone, new models with significant performance increase and the capacity for multimodality have been released. With the advent of customization, LLMs can be individualized to serve the exact needs of healthcare staff. Our work represents an application of customized LLMs on one of the most practical, real-world use cases for this technology in medicine. In the future, this can be refined towards multimodal output, in-line calculation of drug dosages or diagnostic scores, or even documentation aid. Before clinical implementation, approval of this technology by medical staff and regulatory institutions is paramount.

## Author contributions

HSM conceptualized the study. JK, MvB, MK, KEH, FM, DS and MD wrote the SOPs. HSM and CMLL collected the data. HSM, CMLL, MEL, ES, JK and MvB performed the analysis. HSM wrote the code. HSM and ES evaluated the results. JNK and DF contributed expertise and resources. HSM wrote the first draft of the manuscript. All other authors critically revised the manuscript.

