## Supplementary material for "Customizing GPT-4 for clinical information retrieval from standard operating procedures"

| **GPT** | **System prompt** |
| --- | --- |
| SOPHIA | You are an assistant supplied with standard operating procedures (SOPs) of a German university hospital. You answer doctors' questions about the content of SOPs provided by their hospital. The SOPs will be provided in PDF format and can contain text, graphs and images. The questions will be in German and you should answer in German. In your answers, you should strictly adhere to the information provided in the SOPs. The first sentence of every answer should state the name of the SOP. Then, the answers should contain a brief context of the desired information and then a precise answer to the user's question. You must not add any information that is not contained in the given SOP files. Questions that cannot be answered with any of the given files must not be answered at all. If a question has not been asked precisely enough, you are allowed to ask for more detailed instructions. If you find conflicting content in multiple files, say so. |
| CARL | You are a doctor supplied with standard operating procedures (SOPs) of a German university hospital. You answer doctors' questions about the content of SOPs provided by their hospital. The SOPs will be provided in PDF format and can contain text, graphs and images. The questions will be in German and you should answer in German. In your answers, you should strictly adhere to the information provided in the SOPs. The first sentence of every answer should state the name of the SOP. Then, the answers should contain a brief context of the desired information and then a precise answer to the user's question. You must not add any information that is not contained in the given SOP files. Questions that cannot be answered with any of the given files must not be answered at all. If a question has not been asked precisely enough, you are allowed to ask for more detailed instructions. If you find conflicting content in multiple files, say so. |

### Suppl. Table 1

System prompts used to generate SOPHIA and CARL.

| **SOP** | **Degree of detail** | **English translation** |
| --- | --- | --- |
| antibiotic treatment standards | low | What is the antibiotic of choice for suspected septicaemia? |
|  | intermediate | I admitted a patient with moderate pneumonia. Which antibiotic should I use and for how long? |
|  | high | A young female patient with a history of iv drug abuse has tricuspid valve endocarditis. Blood cultures have been taken, the result is not yet available. What is the best calculated antibiotic therapy? |
| pancreatic cancer | low | On which postoperative days do I have to determine amylase in the drainage after a pancreatic head resection? |
|  | intermediate | My patient underwent pancreaticotomy yesterday. When can I start to build up the diet? |
|  | high | My patient with pancreatic carcinoma has undergone pancreatic resection and the anastomosis is assessed as high-risk. How and for how long do the drains need to be flushed? |
| COVID-19 | low | Which Covid-virustatics can I give with GFR <30? |
|  | intermediate | A 64-year-old, fit patient has Covid. Do I have to treat him? |
|  | high | A 60-year-old female patient with non-Hodgkin's lymphoma has Covid with a high O2 requirement. The GFR is 13, what is the treatment of choice? |
| intestinal cleansing | low | What should my patient eat before a colonoscopy? |
|  | intermediate | A patient is coming for a colonoscopy tomorrow and asks for signs of adequate bowel irrigation. What can I tell her? |
|  | high | A patient with suspected colon carcinoma has received 4 sachets of powder in preparation for a colonoscopy and asks on which days he should take which sachets. When should which sachet be taken? |
| neutropenia | low | When do I have to hospitalise a patient with suspected febrile neutropenia? |
|  | intermediate | I have a patient with a fever in neutropenia who I am treating as an outpatient. How many days should I treat her with antibiotics? |
|  | high | I am admitting a patient as an inpatient. The neutrophils are 0.9, the temperature is currently 39.3 and the patient is rather unstable. Which antibiotic should I give him? |
| intraperi- toneal chemothe- rapy | low | Do patients have to be admitted to an intensive care unit after surgery with HIPEC (Hyperthermic intraperitoneal chemotherapy) ? |
|  | intermediate | A patient with gastric carcinoma on my ward had an operation with HIPEC 3 days ago. When can I remove the abdominal Robinson drains? |
|  | high | I have admitted a patient with gastric carcinoma. He will be operated on tomorrow and will then undergo HIPEC. He has reduced kidney function. What should I look out for? |
| opioids | low | Which opioids can I best use for patients with renal insufficiency? |
|  | intermediate | I would like to treat a patient with tilidine/naloxone. What dosage should I start with? |
|  | high | One patient is currently not receiving adequate analgesia with 12 mg hydromorphone orally per day. I would like to change the analgesia. What is the dose equivalent for oxycodone? |
| oesopha- geal cancer | low | What should I look out for after an esophagectomy? |
|  | intermediate | My patient had an esophagectomy 2 days ago. When will he be able to eat again? |
|  | high | A 67-year-old patient with esophageal carcinoma is on my ward following an esophageal resection. The operation was 3 days ago. Today the gastric tube slipped out during mobilization. There was about 500ml per day in the gastric tube. What should I do now? |
| colorectal cancer | low | Which examinations are part of colon CA follow-up care? |
|  | intermediate | A patient undergoing follow-up colon cancer care has come for an ultrasound scan and the sound conditions are extremely poor. Is there a CT indication? |
|  | high | A patient with post-operative colon carcinoma underwent surgery a year ago. Today, an adenoma was found in the follow-up colonoscopy. When do I need to book the next check-up? |
| CAR-T-Cell therapy | low | How do I diagnose an ICANS? |
|  | intermediate | My patient with CRS grade 2 has no symptom improvement after 2 doses of tocilizumab. How do I organize further therapy? |
|  | high | A patient has an ICE score of 2, a fever and is hypotensive. What does she have and how should I treat her? |

### Suppl. Table 2

**Question catalogue**. All questions which were used for evaluation listed with their respective SOP and their respective degree of detail as described in the methods. The original questions were in German, this table shows the English translation.

| **Parameter** | **GPT-4** | **Claude-3-opus** |
| --- | --- | --- |
| embedding model | text-embedding-3-large (OpenAI) | text-embedding-3-large (OpenAI) |
| distance function (embedding) | cosine similarity | cosine similarity |
| chunksize | 1048 | 1048 |
| chunk overlap | 50 | 50 |
| temperature | 0 | 0 |
| model | gpt-4  (OpenAI) | Claude-3-opus-20240229 (Anthropic) |

### Suppl. Table 3

**Hyperparameters**. Hyperparameters which were used for API-based comparison of GPT-4 and Claude-3-opus.
